# *ShockModes:* A Multimodal Model for Prognosticating Intensive Care Outcomes from Physician Notes and Vitals

**DOI:** 10.1101/2022.12.16.22283559

**Authors:** Ridam Pal, Shaswat Patel, Akshala Bhatnagar, Hardik Garg, Pradeep Singh, Ritesh Singh Soun, Aditya Agarwal, Aditya Nagori, Ashish Khanna, Rakesh Lodha, Piyush Mathur, Tavpritesh Sethi

**Affiliations:** Indraprastha Institute of Information Technology Delhi, India; Department of General Anesthesiology, Anesthesiology Institute, Cleveland Clinic, Cleveland, Ohio, USA; Department of Anesthesiology, Section on Critical Care Medicine, Wake Forest School of Medicine, North Carolina, USA; All India Institute of Medical Sciences, New Delhi, India

**Keywords:** Shock Index(SI), Named Entity Recognition(NER), Transformer Architecture, Machine Learning, Natural Language Understanding

## Abstract

**Objective:** Shock Index (SI) is widely used for prognosticating outcomes in ICU and emergency settings. We aimed to create a multi-modal early warning system (EWS) for development of abnormal shock index using routinely available vitals and clinical notes.

**Material and Methods:** 17,294 ICU-stays in MIMIC-III data were scored for SI. A new episode of abnormal SI was defined as SI > 0.7 for >30 minutes AND preceded by >=24 hours of normal SI. ICU stays with <24 hours admission, or SI >0.7 within the first 24 hours of admission, or missing SI in >50% in the 24 hour early warning window were excluded, leaving a final cohort of 337 normal and 84 abnormal SI instances. 3117 features from vitals time-series combined with BERT-based features from clinical notes were used to train a battery of machine learning models. The best multimodal pipeline (*ShockModes*) was assessed for interpretability using SHAP features.

**Results:** Vitals-based, notes-based and multi-modal classifiers achieved the best sensitivity of 0.81, 0.81, and 0.83 with corresponding specificity of 0.92, 0.99, and 0.94 respectively, thus demonstrating the potential of *ShockModes* for early detection, while preventing false alarms. Global SHAP values revealed Fourier-features of heart rate and heparin sodium prophylaxis as top features. Sensitivity of early detection was highest in acute respiratory failure and chronic kidney disease patients.

**Conclusion:** The multimodal, interpretable early warning system *ShockModes* can be used for prognosticating SI based outcomes in ICU and emergency settings.

## INTRODUCTION

Data and predictive models are being used increasingly to augment decision making in Intensive Care Units (ICUs). Early warning signals to predict physiological decompensation in ICU patients can trigger early interventions, improve outcomes and potentially save patient lives in the ICUs [1–4]. The widespread adoption of electronic medical records (EMRs) has accelerated the development of early warning systems by making large volumes of data available, potentially on a real-time basis. Accessibility to EMRs enables clinicians to make use of numerous modalities of information such as patient history, vitals, laboratory investigations, treatment response and other diagnostic modalities to evaluate patient status in the ICU. This assists in designing risk scores such as the Sequential Organ Failure Assessment (SOFA) score for diagnosing sepsis-related organ failure. The scoring criteria often include heterogeneous and non-simultaneous measurement such as vitals, clinical evaluation and laboratory investigations, typically not available simultaneously in real-time. Moreover, most of these scoring criteria require subjective clinical assessment and have changing definitions which make these difficult to be consistently implemented. While sepsis remains one of the most commonly modeled outcomes in the ICUs the nebulous definition of sepsis and the related scoring criteria limits the utility of models that have been developed using a combination of structured, unstructured and vitals data[5–7]. Most of these studies concluded that there is an additional value of including unstructured clinical data to improve the prediction of models[6,7].

Data-driven scoring approaches in the ICUs, on the other hand, allow continuous scoring using real-time vitals that can be combined with daily lab investigations and clinical notes. Shock index (SI) which is defined as the heart rate (HR) divided by systolic blood pressure (SBP) serves as a clinical outcome predicting tool for bedside assessment, risk stratification and prognostication.[8,9] The use of shock index has been examined in predicting outcomes in emergency and low resource settings for various conditions including sepsis, hemodynamic shock, acute myocardial infarctions, stroke, advanced cancer, pulmonary embolism, pneumonia and trauma.[9–11] These outcomes in their early stages are more likely to respond to therapy, whereas once irreversible end-organ damage sets in, chances of mortality is high.[12] Thus, prediction and early identification of deterioration in shock index can support timely life-saving interventions.[13,14]

The utility of Shock Index extends beyond the clinical evaluation of Shock, although that is the most frequently modeled use case. Previously, thermal imaging[15] and time series data from the vitals[16] has been used to predict abnormal shock index across age, ICU specialties and geographies. Similarly, a multivariate combination of vitals was used to create machine learning models for shock prediction.[17–19] However, none of these studies has addressed Shock Index using a multi-modal approach. The current study addresses this gap and combines structured and unstructured data present in EMRs for prognosticating deterioration in the Shock Index. We hypothesized that (i) physician notes encapsulate the sum total of underlying diseases (e.g., diabetes), medications, and other indicators of infection, e.g. fever, use of antibiotics, etc. These information can be mined into features using representation learning with the help of language models. (ii) The strong association of vital signals, such as heart rate and BP with Shock index makes this an important feature for abnormal shock prediction.

The objectives of this study were (i) Model Shock Index as an objective indicator of outcomes, (ii) Assessment of improvement, if any, in model performance and clinical interpretability of the multi-modal models as compared to vitals or physician notes alone, (iii) Efficacy of model performance for diagnosing diseases across clinically segregated cohorts. To this end, we created *ShockModes*, an end-to-end multi-modal pipeline by fusing vitals signals with medications and contextual information such as history of present illness (HOPI), for 24-hour ahead prediction of abnormal shock index (Figure 1).

**Figure 1:**
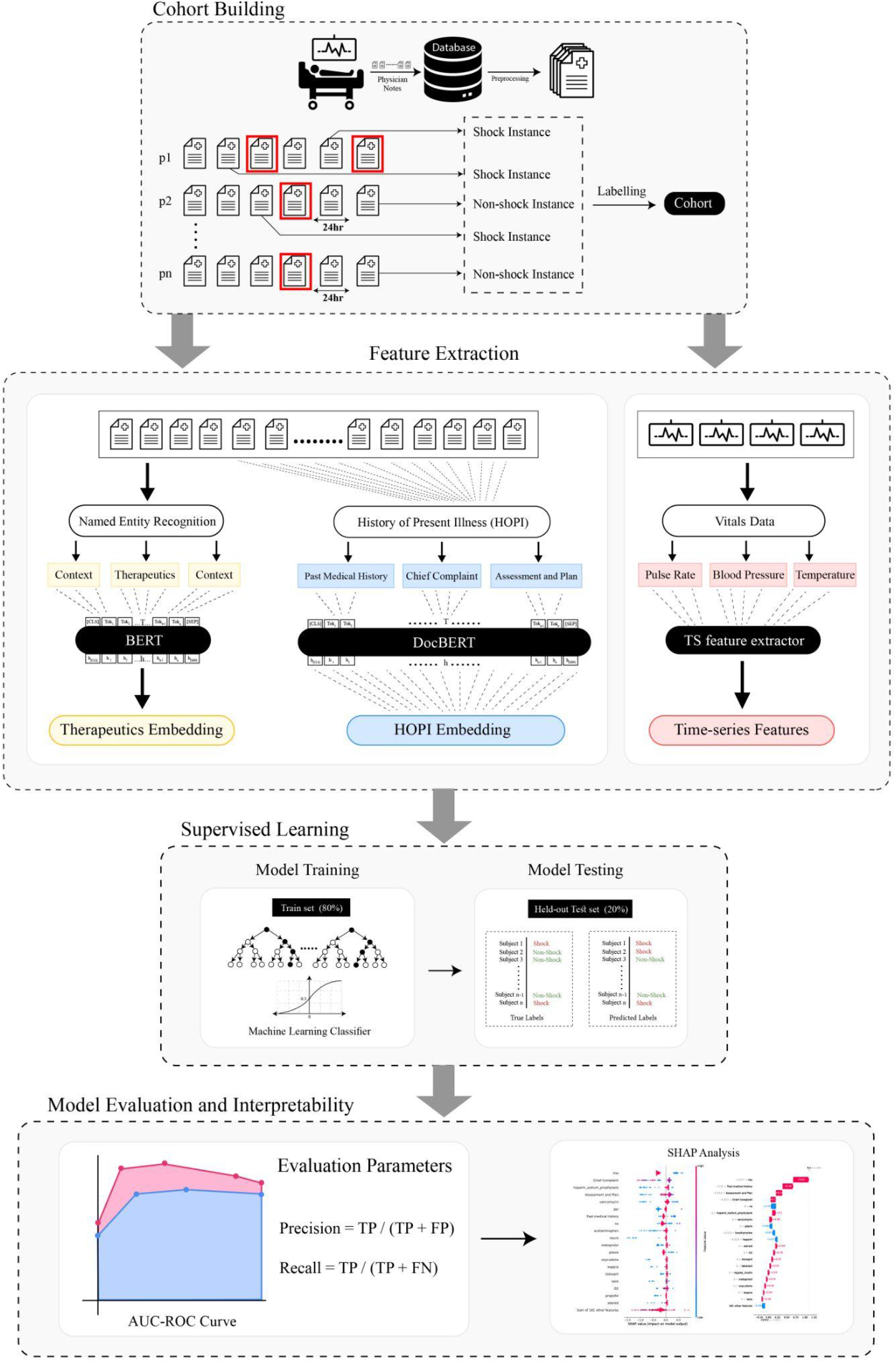
Overview of ShockModes pipeline. ShockModes takes physician notes and vitals data as input, clinical entities (Therapeutics & History of Patient Illness) are extracted using MED7 Named Entity Extraction(NER) whereas Time-Series (TS) features are extracted using tsfresh python module. Unsupervised Embeddings fused with TS features serve as input features for the machine learning models for predicting the onset of abnormal SI with 24-hours lead time. SHAP analysis reveals interpretable features for global and patient-specific notes.

## METHODS

### Dataset

Our study has been conducted using the publicly available Medical Information Mart for Intensive Care (MIMIC) III v.1.4 database.[20] The MIMIC-III database provided critical care data for over 40,000 patients admitted to intensive care units at the Beth Israel Deaconess Medical Center (BIDMC). The Noteevents data contained textual information of physician notes corresponding to different categories. We curated this clinical database to create 24-hour patient encounter cohorts of vitals and text data from physician notes. We have also extracted the patients’ heart rate (HR), systolic blood pressure (SBP), respiratory rate (RR), and oxygen saturation (SpO_2_) for every 24 hours recorded in the critical care setting.

### Cohort Construction

Shock index was defined as the heart rate (HR) divided by systolic blood pressure (SBP), and has been used for assessment of hemodynamic instability in suspected shock patients. Continuous vitals data corresponding to each patient was summarized at 1-minute resolutions and utilized to calculate the SI. Epochs of 30-minute length were generated. The onset time of abnormal shock index was defined as the starting time of an epoch in which the median shock index value was greater than 0.7. The corresponding abnormal shock index labels were calculated using a cut-off value of 0.7 for the shock index (SI ≥ 0.7 corresponds to abnormal shock index, SI<0.7 corresponds to normal shock index and NA corresponds to missing label).[11,14] Days for which the final label was missing were discarded. A new instance of abnormal SI was defined as a >30 minutes length episode of abnormal SI preceded by at least 24 hours of normal SI. Only new instances of abnormal SI were included in the cohort as ongoing abnormal SI may be a trivial predictor for future abnormal SI. For text, we have considered only the top seven frequencies of physician note types. The seven notes from the physician category included notes from physician residents, intensivists, the physician attending, and other ICU notes which encompassed the majority of the clinical data that could be extracted from the physician notes. Physician notes were mapped using date, subject ID and ICU stay ID to their next-day labels generated using continuous vitals data. Our cohort exclusively for textual data comprised 355 instances of normal SI patients and 87 abnormal SI patients (Supplementary Table 1). Our vitals data and multi-modal cohort comprised 337 normal instances and 84 abnormal SI patients. Table 1 summarizes the patient characteristics for the extracted multimodal cohort.

**Table 1.** Multimodal Cohort characteristics. Values are median (IQR) unless indicated, *signifies Wilcoxon (W) Ranksum test (non-parametric) or Student’s t-test (t) (parametric) which were used after testing for the normality assumption. c - Chi-squared test of proportions.

| Variables | Median (IQR) |  | Statistical Test |
| --- | --- | --- | --- |
|  | Normal Shock Index<br>(n=334) | Abnormal Shock Index<br>(n=87) | p-value |
| Age | 69.87 (21.97) | 68.99 (18.67) | 0.943 (w)* |
| Length of stay(LOS) | 5.16 (11.26) | 4.40 (8.77) | 0.183 (w)* |
| Heart Rate, per min | 67.31 (10.00) | 72 (10.25) | $5.82 \times 10^{-07}$ (w)* |
| Pulse Rate, per min | 67 (11.00) | 71 (10.25) | $4.41 \times 10^{-06}$ (w)* |
| Respiratory rate,<br>per min | 18 (4.94) | 18.75 (4.00) | 0.119 (w)* |
| Oxygen Saturation | 97 (3.00) | 97 (3.63) | 0.899 (w)* |
| Gender (Female %) | 38.28% | 38.1% | 0.999 (c)* |

### *ShockModes* Pipeline

#### Preprocessing

Text formatting was initially done to extract the therapeutics from physician notes using Named Entity Recognition(NER), details of which are provided in (Supplementary Figure 1).[21] The encrypted information ([**word**]) along with single-letter words (example F, M etc), present in the MIMIC data were removed from the text. Text was converted to lowercase followed by standard pre-processing such as removal of whitespaces, stopwords, punctuations, digits, and words with less than or equal to two letters. The misspelled words were rectified using Levenshtein distance and the resulting vocabulary was also vetted by a clinician. While training the language model, the numeric values were considered for minimizing the loss of contextual information present in the form of dosage, frequency, and other critical pieces of information.

### Feature Extraction

Named Entity Recognition was performed using MED7[22] to extract therapeutics from physician notes. Therapeutics consisting of more than one word were concatenated with an underscore character.(e.g. metoprolol tartrate => metoprolol_tartrate).

#### Word2Vec and Transformer based Therapeutics embeddings

A low-dimensional representation (100 dimensions) for the therapeutics present in the corpus of the physician notes was learned using the Word2Vec model with skip-gram algorithm, one-hot encoding, and a fixed window size of five using the Gensim library.[23] For Transformer-based models, WordPiece tokenization[24] was carried out to to avoid out-of-vocabulary issues. BioBERT[25] and BioClinical BERT[26] were then tuned using a sliding window of 512 tokens/subwords with the therapeutic entity in the middle of the window. The resulting embeddings for BioClinical BERT and BioBERT were 768 and 1024 dimensional, respectively. Since each word may have multiple embedding vectors based on their context within the document, for each document a mean embedding vector for each therapeutic was generated.

#### History of Present Illness (HOPI) Embeddings

To incorporate clinical context we considered i) Chief complaint, ii) Assessment and plan, and iii) Patient history, together known as History of Present illness(HOPI). The chief complaint section constituted the present issues of the patient with suspicion of shock, time of onset, duration, and the progression of the clinic sign & symptoms in detail. It also includes previous vitals and initial management done(if available) at the time of evaluation during admission. Assessment & plan comprises the provisional management scheduled based on the initial clinical evaluation. Past medical history along with the HOPI contributes to the initial management by identifying risk factors, etiological causes and presenting stages of shock. Each component contains sequence tokens which can potentially exceed the input sequence constraint of 512 tokens hence models that are capable of addressing this challenge were explored.[27–29]. Low-dimensional representation of HOPI components was learnt using Doc2vec[30] with distributed memory algorithms, and the other parameters have been supplemented(Supplementary Table 2).

For the Transformer based approach, a DocBERT[29] was utilized for extracting low-level representation for each of the above-mentioned components. The entire textual input was split into multiple chunks and provided to the model for feature extraction. All the [CLS] tokens from each chunk were aggregated to obtain a vector representing the complete textual input.

A hybrid architecture that combined embeddings generated from medications and HOPI was developed in which the embeddings were fused together to serve as inputs for classification models. A comparative analysis related to token lengths and subword length for BioBERT and BioClinical BERT has been supplemented (Supplementary Figure 2).

### Vital Features

A collection of 3117 time series features (TS features) was extracted using the “tsfresh” Python module, which includes linear and nonlinear physiological characteristics such as autocorrelation, sample entropy, linear trends, statistics, etc.[31]

### Model Development and Validation

The cohort was randomly sampled into a stratified training test split in the ratio of 80:20. All the patient ids with multiple instances were present either in the training or testing dataset. Each drug in the corpus along with HOPI has been considered as a feature, where the mean of these embeddings has been treated as the feature value for the classifier. Words directly indicative of shock were masked. Vitals features combined with mean embedding of therapeutics and HOPI have been considered as input features for *ShockModes*. We had an extensive set of features for the classifier, which would have led to the overfitting of the models and increased the time complexity. In order to reduce features, feature selection was performed using Extra Tree classifiers[32] present in the scikit-learn library.[33] Since the dataset was highly imbalanced, we have performed oversampling using SMOTE[34] present in the scikit-learn library. Machine learning models including Logistic Regression, Random Forest, GradientBoost, AdaBoost, and XgBoost[35–39] were trained on the vital time series features combined with learned embeddings of therapeutics and HOPI for predicting the onset of the abnormal SI. While implementing the algorithms in the training dataset, we have initialized most of the parameters with the default value, and hyper-tuned some features based on our task. The algorithms were tuned to have higher sensitivity to be used as a screening approach. The tunable parameters for the models are listed in Supplementary Information(Supplementary Table 2). Model performance indicators were evaluated using bootstrap sampling for 100 iterations to record the mean value. AUC-ROC score and F1-scores were used to select the best pipeline from each category.

### SHAP Analysis for Interpretability

Shapley Additive Explanations (SHAP) utilize Shapley values to calculate feature importance while explaining the model’s output and providing an interpretability interface.[40]

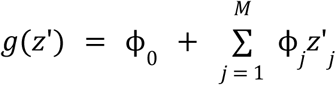

Where g is the classifier, *z*′ is the simplified features of therapeutics and HOPI, M is the maximum number of simplified features of therapeutics and HOPI, and ϕ _*j*_ ∈ *R* is the Shapley values associated with feature j. The training dataset has acted as prior knowledge from which prior probability is calculated, the model’s output probability can then be explained with respect to the prior probability. Specific instances of patients were explained using waterfall plots for depicting the most important features which lead to the prediction of abnormal/normal SI. Global feature importance for the entire dataset has been visualized using the bar plots and beeswarm plots. The Shapley values were calculated for therapeutics and HOPI and provide a window into the inner workings of the classifiers used for predictions.

### Implementation of Pipeline

We have cited the software and algorithms that have been used for developing the novel architecture (Supplementary Table 3). Bi-directional transformer models (BioBERT, BioClinical BERT) have been implemented using the Hugging Face library[41]. The models were trained using Tesla P100-SXM2 with 512GB RAM, and 16GB Nvidia GPU.

## RESULTS

### Exploratory Data Analysis

The multimodal cohort was constructed from 17,294 ICU-stays in MIMIC-III data comprising 334 normal and 87 abnormal instances of SI. The median of heart rate and respiratory rate for abnormal SI was recorded as 72/min and 18.75/min respectively, greater than the values recorded for normal SI. Other characteristics of the cohort were reported in Table1.

The textual modality was constructed from 2,083,180 clinical notes in MIMIC-III, the categories with the top seven frequencies in the physician notes (141,624) were taken to understand the context of the notes (Figure 2). Among the Physician notes, physician resident progress note (62,682) was the most common category followed by intensivist notes. A WordCloud and frequency plots of most(N=20) common therapeutics shows antibiotics, heparin, lasix/furosemide among the most frequently occurring in the data.

**Figure 2:**
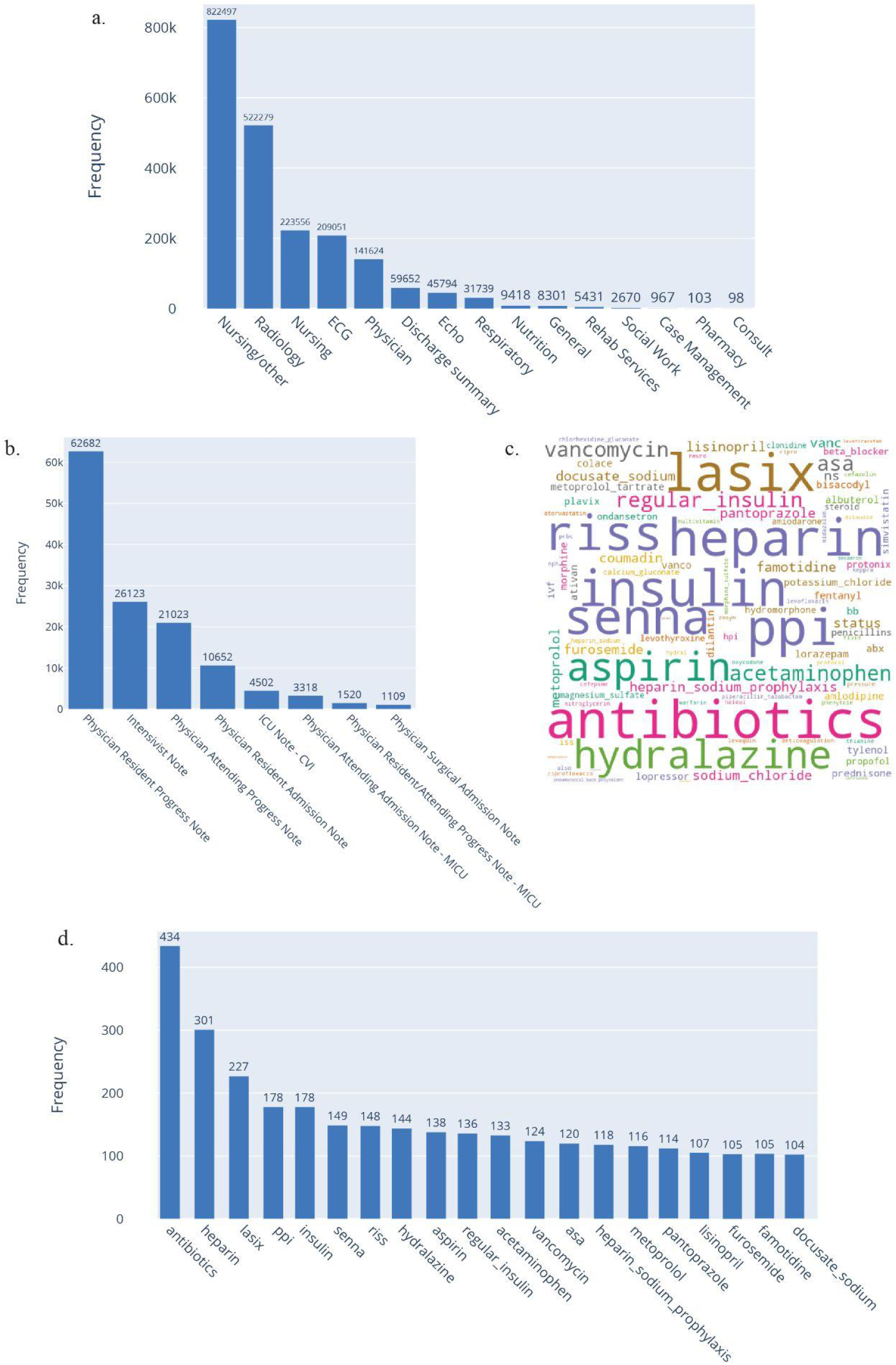
a) Plots illustrating the frequency of different categories of treatment notes. b) Frequency of Physician note types. c) Word Cloud of Therapeutics present in the corpus. d) Top-20 most common therapeutics encountered during periods with abnormal SI.

### Embedding features and model performance

The model performance indicators for the variety of language models developed indicate that there was no single combination that outperformed all the others on all metrics. The transformer based BioBERT and DocBERT embeddings combined with Gradient Boosted model (learning rate of 0.1, n_estimators of 100, maximum depth of 3 and minimum samples split of 2) achieved the best performance based on combined AUC-ROC and F1-score of 0.68 and 0.76 respectively. Taking sensitivity as a metric for screening, Adaptive Boosting trained on Word2Vec and Doc2Vec achieved the highest sensitivity of 81% with F1-score of 74%. While taking specificity as screening metric, RandomRandom Forest trained on BioBERT and DocBERT achieved the highest specificity (0.99). This depicts that simpler embeddings capture the context within the history of present illness and medications and performed comparably to transformer models. We believe that this may be because of the preliminary step of NER which simplifies the complex task instead of working with the raw text. Model parameters and model performance indicators for all combinations are described in Table 2.

**Table 2.**
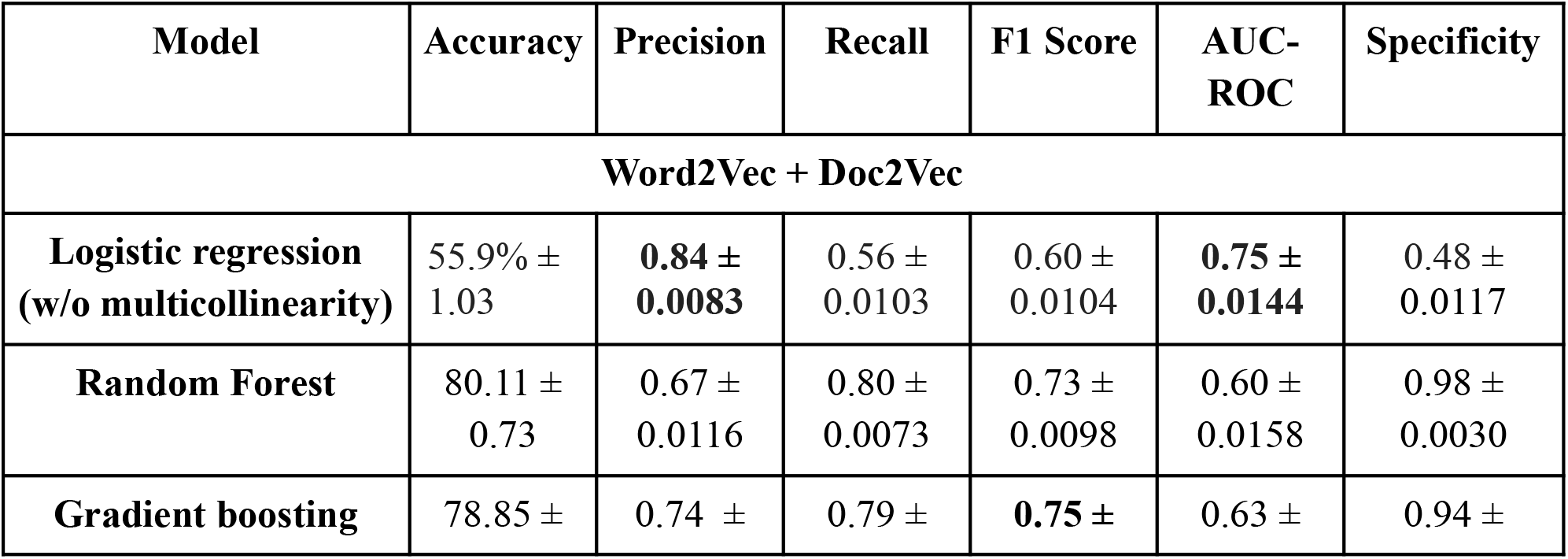
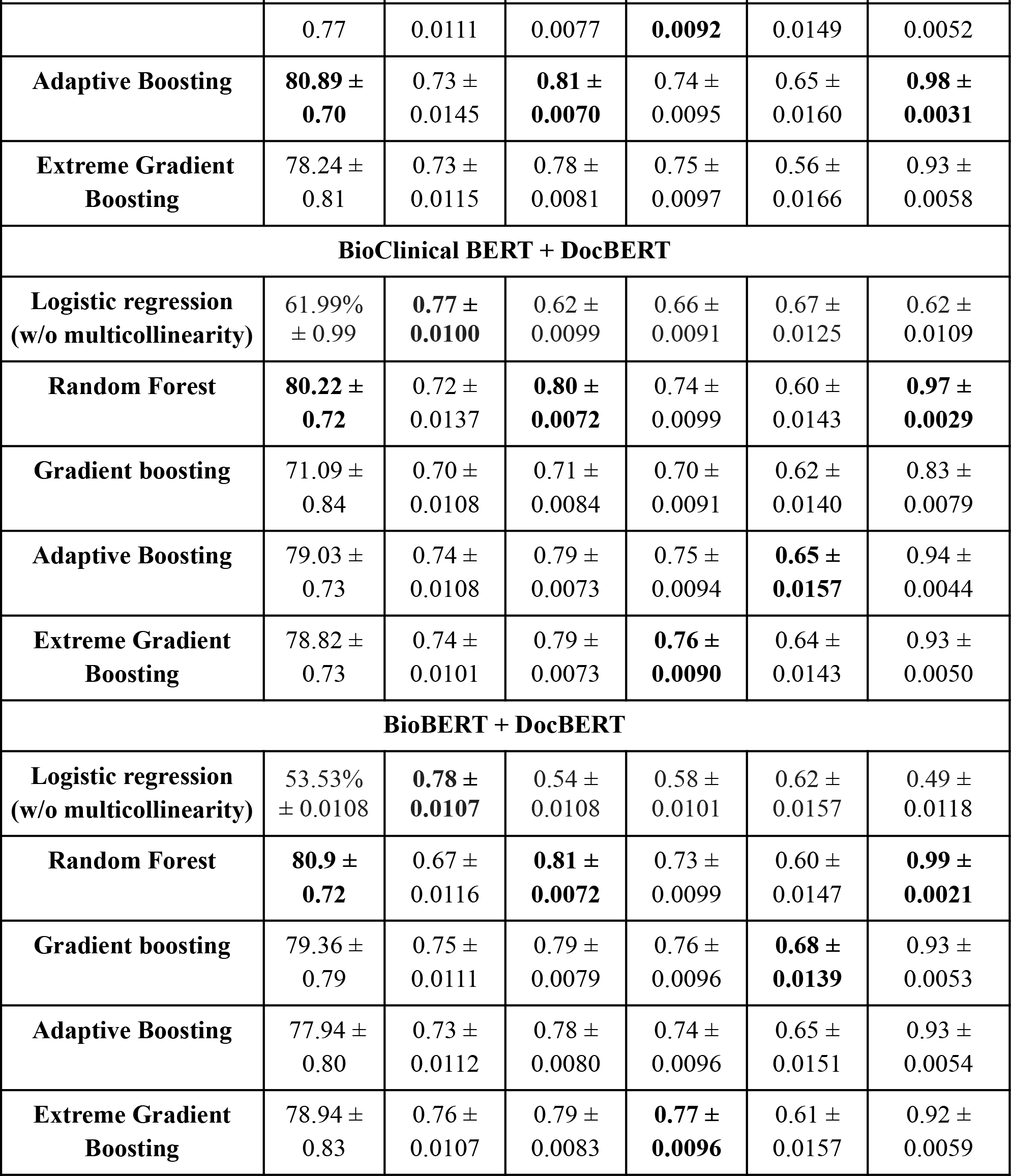
Using the clinical notes features alone, results of SI (abnormality) prediction on a 24-hour cohort of MIMIC III dataset with a margin error of one standard-deviation from the mean. Random Sampling(N=100) of the test dataset, with Bootstrap iterations of 100 has been recorded for the mean value. Only Therapeutics and HOPI embeddings were considered as input features for the Machine learning models. We considered the Gradient Boosting model with BioBERT+DocBERT embedding as the best model since it recorded a high F1-score and AUC-ROC score.

### Leverage gained from History of Present illness and length of context

Since HOPI is expected to capture the patient’s trajectory, an ablation experiment to understand the importance of the HOPI in the Physician notes was carried out (Figure 3a,3b). We observe a significant increase (p-values ≤ 0.05) in AUC-ROC score with the incorporation of HOPI embeddings for three models (Supplementary Table 4). Comparison among the models (with/without HOPI as feature) is shown in Supplementary Table 5. Therefore, we find that the context encapsulated in HOPI is important for performance of models predicting the outcomes in patients admitted to the ICU and must be included. We have also empirically vetted the fallout in performance of 768 sized embeddings for HOPI against its mean embeddings (Supplementary Table 6). For Transformer based models, we hypothesized that the length of context (window length) would be positively correlated with model performance indicators (Figure 3c).

**Figure 3.**
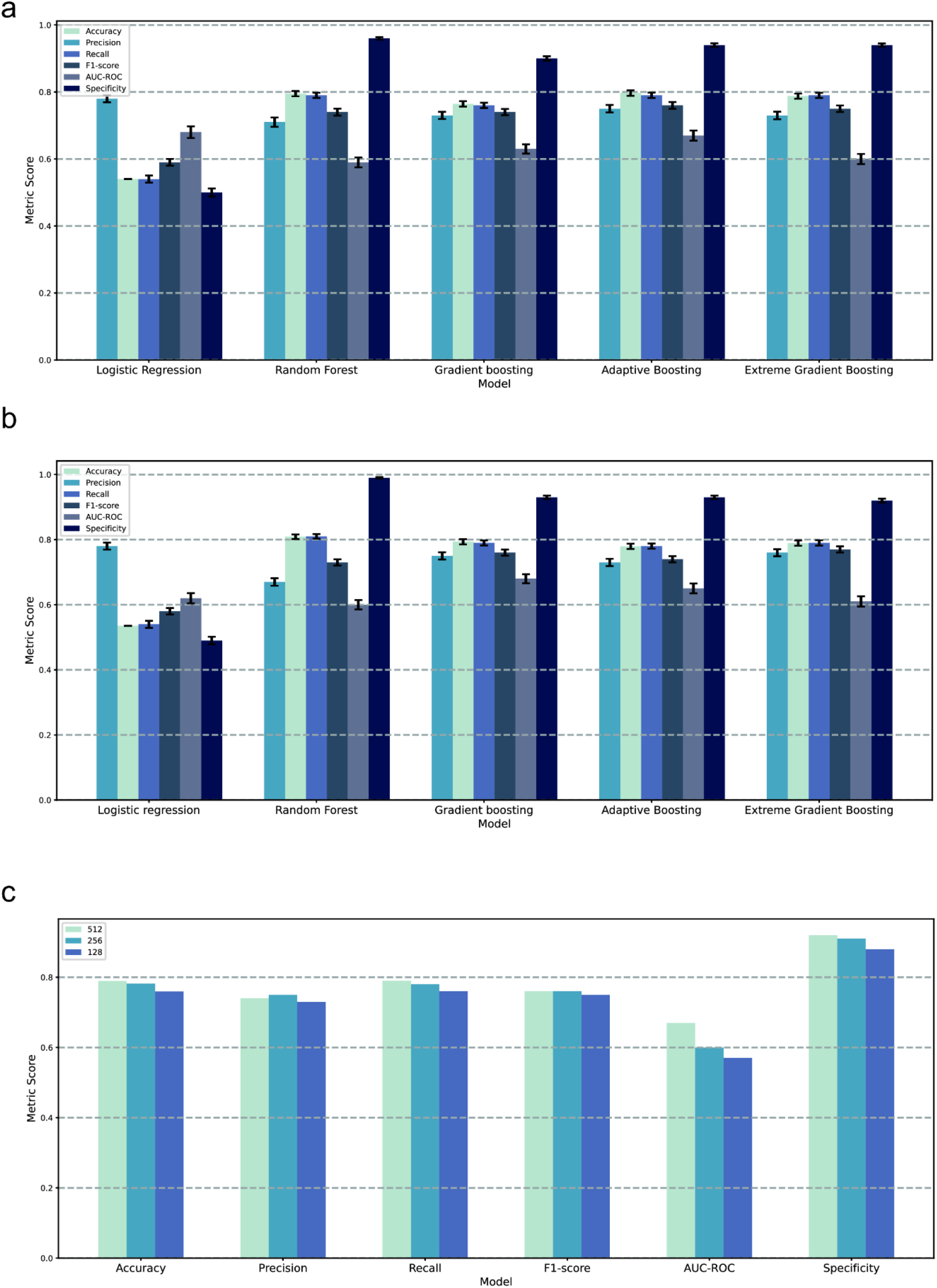
Leverage gained with addition of History of Present illness and length of context. Efficacy of various machine learning models a) exclusive and b) inclusive of HOPI as features. Therapeutic embeddings were generated from BioBERT while HOPI embeddings were generated from DocBERT. c) Comparison of different contextual window lengths (512, 256, 128) for the embeddings generated from BioBERT. These embeddings were compared against different metrics for the best pipeline exclusively of textual data(Gradient Boosting+DocBERT embedding+BioBERT contextual embedding). The plot depicts that sequence length of 512 has achieved the best AUC-ROC demonstrating that with an increase in the sequence length, there has been an increment in the AUC-ROC.

### Analysis using only Vitals Data

We trained a battery of machine learning models exclusively on top 200 TS features processed from vitals data to verify its performance over clinical notes (Table 3). Considering both AUC-ROC and F1-score as screening parameters we observed that vitals data alone outperforms textual embeddings data by 6% in terms of AUC-ROC. Adaptive Boosting was the best model for vitals data, as compared to Gradient Boosting model having BioBERT+DocBERT embeddings as features. Notably, Random Forest achieved the best specificity and sensitivity of 0.92 and 0.81 respectively.

**Table 3.** Results of SI (abnormality) prediction exclusively using vitals data on a 24-hour cohort of MIMIC III dataset with a margin error of one standard-deviation from the mean. Only TS-features extracted from vitals data were considered as input features for the Machine learning models. Random Sampling(N=100) of the test dataset, with Bootstrap iterations of 100 has been recorded for the mean value.

| Model | Accuracy | Precision | Recall | F1 Score | AUC-ROC | Specificity |
| --- | --- | --- | --- | --- | --- | --- |
| **Logistic Regression (w/o multicollinearity)** | 0.60 ± 0.0093 | 0.67 ± 0.0110 | 0.60 ± 0.0093 | 0.63 ± 0.0092 | 0.42 ± 0.0128 | 0.69 ± 0.0100 |
| **Random Forest** | **0.81 ± 0.0076** | **0.80 ± 0.0087** | **0.81 ± 0.0076** | **0.80 ± 0.0080** | 0.73 ± 0.0122 | **0.92 ± 0.0066** |
| **Gradient boosting** | 0.72 ± 0.0078 | 0.73 ± 0.0088 | 0.72 ± 0.0078 | 0.72 ± 0.0078 | 0.59 ± 0.0132 | 0.82 ± 0.0079 |
| **Adaptive Boosting** | 0.81 ± 0.0077 | 0.79 ± 0.0090 | 0.81 ± 0.0077 | 0.79 ± 0.0083 | **0.74 ± 0.0122** | 0.91 ± 0.0064 |
| **Extreme Gradient Boosting** | 0.76 ± 0.0074 | 0.74 ± 0.0092 | 0.76 ± 0.0074 | 0.75 ± 0.0088 | 0.60 ± 0.0148 | 0.88 ± 0.0067 |

### Multimodal Analysis inclusive of Structured (Vitals) and Unstructured (Textual) Data

Embeddings from therapeutics and HOPI were combined with TS features from vitals data to effectively understand the significance of multimodal features over individuals. We observe that Random Forest trained on Word2Vec and Doc2Vec embeddings inclusive of vitals data achieved an AUC-ROC, F1-score, and Specificity of 0.76, 0.81, and 0.94 respectively outperforming only textual and only vitals based pipelines (Table 4). Taking sensitivity as screening parameter, Random Forest trained on BioBERT and DocBERT achieved the highest sensitivity of 0.83. The high specificity of the models is important to prevent alarm fatigue in the ICU settings, hence emphasized. In general, all the best performing models have a high specificity, thus indicating suitability for sensitive detection, yet limiting alarm fatigue in the early warning settings. This indicates models can leverage further information when both structured and unstructured data are fused together for shock prediction.

**Table 4.** Results of SI (abnormality) prediction combining textual and vitals data on a 24-hour cohort of MIMIC III dataset with a margin error of one standard-deviation from the mean. Random Sampling(N=100) of the test dataset, with Bootstrap iterations of 100 has been recorded for the mean value.

| Model | Accuracy | Precision | Recall | F1 Score | AUC-ROC | Specificity |
| --- | --- | --- | --- | --- | --- | --- |
| **Word2Vec + Doc2Vec** |  |  |  |  |  |  |
| **Logistic Regression (w/o multicollinearity)** | 0.60 ± 0.0093 | 0.67 ± 0.0110 | 0.60 ± 0.0093 | 0.63 ± 0.0092 | 0.41 ± 0.0127 | 0.69 ± 0.0100 |
| **Random Forest** | 0.83 ± 0.0068 | 0.81 ± 0.0092 | 0.83 ± 0.0068 | 0.81 ± 0.0081 | **0.76 ± 0.0116** | **0.94 ± 0.0050** |
| **Gradient boosting** | 0.74 ± 0.0072 | 0.74 ± 0.0085 | 0.74 ± 0.0072 | 0.74 ± 0.0074 | 0.60 ± 0.0144 | 0.84 ± 0.0071 |
| **Adaptive Boosting** | 0.82 ± 0.0074 | **0.81 ± 0.0085** | 0.82 ± 0.0074 | 0.81 ± 0.0079 | 0.71 ± 0.0143 | 0.92 ± 0.0061 |
| **Extreme Gradient Boosting** | 0.76 ± 0.0078 | 0.75 ± 0.0090 | 0.76 ± 0.0078 | 0.75 ± 0.0081 | 0.56 ± 0.0151 | 0.87 ± 0.0075 |
| **BioClinical BERT + DocBERT** |  |  |  |  |  |  |
| **Logistic Regression (w/o multicollinearity)** | 0.60 ± 0.0093 | 0.67 ± 0.0110 | 0.60 ± 0.0093 | 0.63 ± 0.0092 | 0.41 ± 0.0127 | 0.69 ± 0.0100 |
| **Random Forest** | 0.79 ± 0.0073 | 0.76 ± 0.0091 | 0.79 ± 0.0073 | 0.77 ± 0.0081 | 0.74 ± 0.0119 | 0.92 ± 0.0065 |
| **Gradient boosting** | 0.75 ± 0.0079 | 0.74 ± 0.0094 | 0.75 ± 0.0079 | 0.74 ± 0.0084 | 0.66 ± 0.0122 | 0.85 ± 0.0071 |
| **Adaptive Boosting** | 0.79 ± 0.0074 | 0.77 ± 0.0086 | 0.79 ± 0.0074 | 0.78 ± 0.0079 | 0.74 ± 0.0128 | 0.90 ± 0.0066 |
| **Extreme Gradient Boosting** | 0.76 ± 0.0076 | 0.75 ± 0.0087 | 0.76 ± 0.0076 | 0.75 ± 0.0078 | 0.63 ± 0.0136 | 0.87 ± 0.0073 |
| **BioBERT + DocBERT** |  |  |  |  |  |  |
| **Logistic Regression (w/o multicollinearity)** | 0.60 ± 0.0093 | 0.67 ± 0.0110 | 0.60 ± 0.0093 | 0.63 ± 0.0092 | 0.42 ± 0.0128 | 0.69 ± 0.0100 |
| **Random Forest** | **0.83 ± 0.0065** | 0.81 ± 0.0088 | **0.83 ± 0.0065** | **0.81 ± 0.0077** | 0.72 ± 0.0147 | **0.94 ± 0.0050** |
| **Gradient boosting** | 0.74 ± 0.0077 | 0.73 ± 0.0095 | 0.74 ± 0.0077 | 0.73 ± 0.0082 | 0.57 ± 0.0128 | 0.85 ± 0.0072 |
| **Adaptive Boosting** | 0.78 ± 0.0077 | 0.76 ± 0.0089 | 0.78 ± 0.0077 | 0.77 ± 0.0082 | 0.71 ± 0.0118 | 0.90 ± 0.0068 |
| **Extreme Gradient Boosting** | 0.75 ± 0.0073 | 0.72 ± 0.0090 | 0.75 ± 0.0073 | 0.73 ± 0.0079 | 0.64 ± 0.0110 | 0.88 ± 0.0068 |

### Model performance across clinically segregated categories

We have analyzed the predictive power of *ShockModes* (multimodal pipeline) by clinically segregating the cohort based on clinical categories (Figure 5). Notably, the best pipeline for *ShockModes* (Random Forest+Word2Vec+Doc2Vec+vitals time-series) was screened on the basis of F1-score and AUC-ROC, whose efficacy was studied across a broad range of disease-related outcomes. *ShockModes* achieved the highest sensitivity (93%) and F1-score (91%) across acute respiratory failure, followed by Chronic kidney disease (92% and 90%, respectively). It achieved better specificity for instances with acute respiratory failure and kidney-related terms (chronic kidney disease), while comparing against (p-value ≤ 0.05) terms suggestive of congestive heart failure. This demonstrates the broad applicability of *ShockModes* across critical care illnesses. We have also analyzed the efficacy of text-based models by segregating the instances of patients based on drugs present in physician notes. Lasix achieved the best F1-score of 0.84 with a sensitivity of 0.89 respectively.

### Interpretability of model predictions using Shapley plots

Figure 4 shows Shapley plots that have been used to analyze the explainability of the models. For interpretability analysis, Gradient Boosting model trained on BioBERT and DocBERT embeddings (textual pipeline) and Random Forest model trained on clinical notes and vital features (multimodal pipeline) were chosen as these performed the best in terms of F1 score and AUC-ROC. The interpretability analysis for all pipelines was supplemented (Supplementary Figure 3, Supplementary Figure 4).

**Figure 4.**
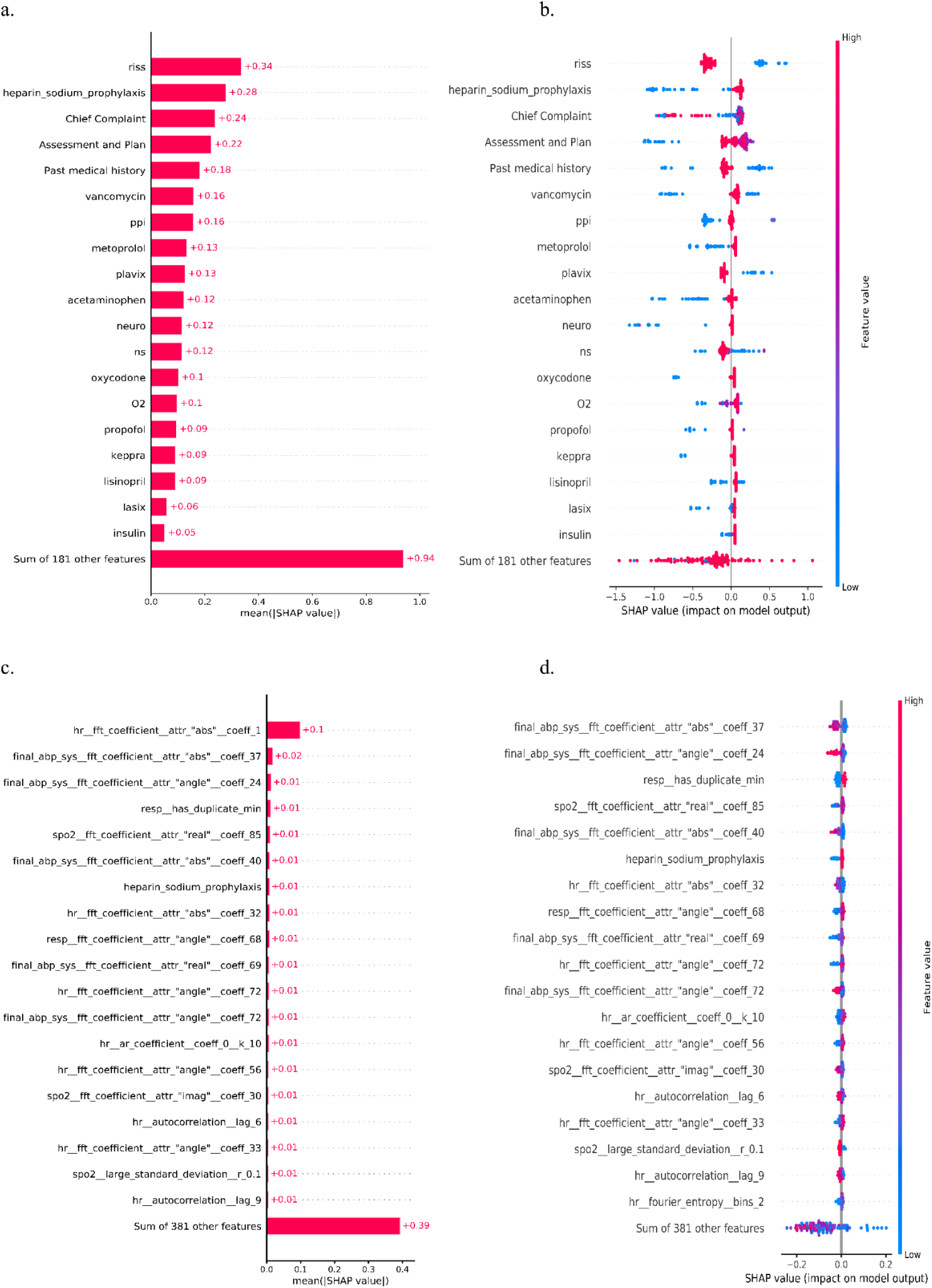
Interpretability of model prediction using SHAP analysis. Sections such as chief complaints, past medical history, and treatment plan along with specific therapeutics such as anticoagulants and antibiotics contain important information for predicting abnormal SI in global feature importance analysis (a) as well as understanding the influence each feature has on the outcome for the textual pipeline. (c) Vitals features were predominant in determining the abnormal SI for the multimodal pipeline as well as certain therapeutics such as Heparin Sodium Prophylaxis turned out to be significant in global feature importance analysis. (b,d) An example of an individual level feature importance for a record is shown as a waterfall plot for textual and multimodal pipeline respectively.

**Figure 5:**
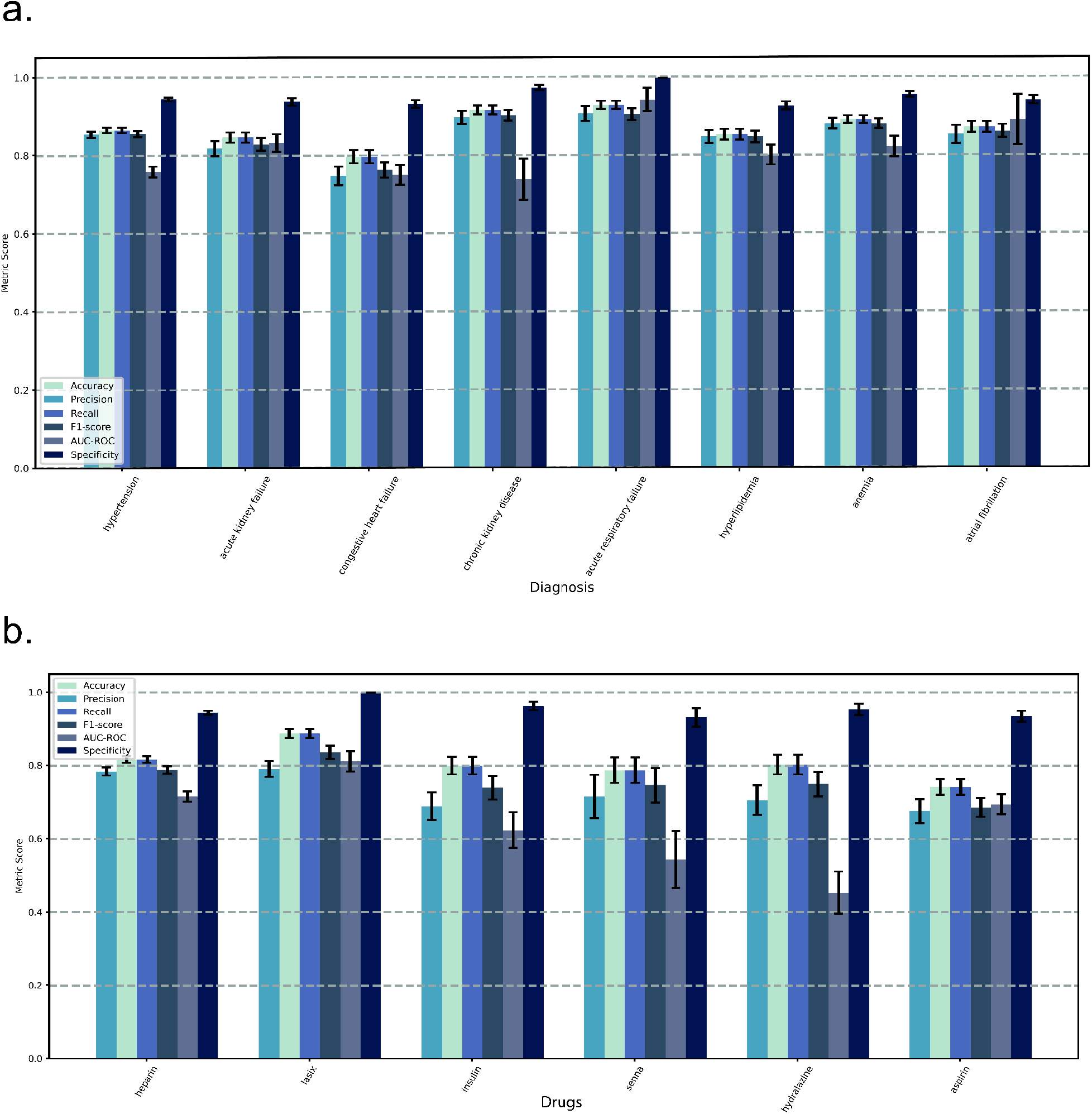
Evaluation of models based on clinical segregation. a) Efficacy of ShockModes on top diagnosis. b) Efficacy of only text-based pipelines on top drugs.

Higher absolute Shapley values are indicative of higher importance. The global feature importance is an aggregate indicator of overall importance of the word across all clinical notes. Figure 4a, 4c depict the global feature importance in the test dataset for textual and multimodal pipelines respectively. RISS (Regular Insulin sliding scale), Heparin sodium prophylaxis and Chief complaints sections were seen as the top ranking global features for predicting worsening of shock index in the textual pipeline. For the multimodal pipeline, SHAP analysis illustrated that most of the top predictors were Fourier-features of heart rate, which is a primary indicator of worsening SI. However, Heparin Sodium Prophylaxis still remained a significant feature for this pipeline, thus leveraging the information contained in clinical notes to improve model predictions when combined with vitals data. Figure 4b, 4d illustrates the impact of these features on the model’s output. It can be observed from the graph that the presence of RISS in a patient’s treatment chart increases the likelihood of downstream development of abnormal SI while the absence of such increases the likelihood of normal SI.

## DISCUSSION

In this study, we demonstrate *ShockModes*, our novel pipeline which exploits the untapped potential of multimodal data for prognosticating 24-hours before the onset of abnormal shock-index. We have shown the effectiveness of our proposed pipeline that utilizes representations learned by various state-of-the-art language models (BioBERT, ClinicalBERT) and vitals data combined with a battery of machine learning models. *ShockModes*, our multimodal pipeline, surpassed individual modalities (clinical notes and vitals data) by nearly 2% in terms of key metrics like AUC-ROC score. Hence, *ShockModes* can serve as an early warning system to make timely interventions, while assisting and accelerating clinicians in an efficient decision making process.

Health condition of patients with abnormal shock index can deteriorate with every minute, hence its diagnosis at an early stage is very crucial. Previous studies have highlighted the challenges in Abnormal shock prediction.[42] Consistent with prior literature, we found methods which utilize both structured (vitals) and unstructured (clinical notes) data for predicting shock-index based outcomes, which also includes shock in ICU.[6,7,43,44] A novel algorithm SERA was proposed which utilized both structured and unstructured data for early prediction of sepsis, while achieving an AUC 0.9 on 24-hours cohort.[7] The use of ClinicalBERT for extraction of meaningful features alongside structured data has shown significant improvements in the early prediction of sepsis shock 4-hour led time.[6] In comparison with their model which used only unstructured data for prediction sepsis, we achieved an AUC-ROC of 0.75 while predicting the outcomes with 24-hours lead time. While combining clinical notes with vitals data, we achieved an AUC-ROC of 0.76. With paucity of available benchmarks, our paper provides baseline models for predicting onset of abnormal SI on a 24-hours cohort.

In Early Warning Systems, higher specificity and sensitivity of prediction is an important component for designing screening systems of ICUs to prevent alarm fatigue due to false positives and false negative cases. This can substantially help in reducing the overhead charges for medical organizations. Drawing conclusions based on specificity and sensitivity values can optimize medical resources as clinicians would otherwise need to perform additional follow-up examination and treatment. Further, a high sensitivity in a clinical setting is a strong indicator of the robustness of the system as it correctly identifies abnormal SI, whereas a high specificity assists in detecting patients with normal SI. Our vitals-based, notes-based and multi-modal pipeline achieved very high specificity values (above 90% for all three cases) indicating the probability of false positive cases to be minimal. This leads to conservation of crucial resources which can be deployed more sensibly in treatment of patients, thereby reducing mortality. The sensitivity also touched 80% in the best case for all three versions of the pipeline. This indicates that a vast proportion of shock patients, who require immediate medical attention, were correctly classified by the model. Hence, this supplements and accelerates the decision making for clinicians, enabling them to make timely interventions and overcoming alert fatigue. Furthermore, *ShockModes* also achieved high sensitivity and specificity values across clinically segregated cohorts, demonstrating the robustness and clinical adaptability of our pipeline. Interpretability of models in clinical settings is very important as it delivers a clear understanding on how the decisions have been made based on input features. Shapely determined Fourier-features of heart rate and heparin sodium prophylaxis as clinically meaningful features of ShockModes for predicting abnormal SI; elucidating the significance of heart rate on our development cohort.

There are few limitations to our current pipeline. Firstly, our pipeline has been exclusively trained on the MIMIC-III dataset which is constrained to adult ICU of specific regions. However, the resulting pipelines can be fine-tuned to other datasets arising from critical care settings for wider adaptability and robustness. Secondly, for interpretability purposes, embeddings for each drug were supplied to the machine learning model which led to sparsity in the matrix. Although this poses the danger of overfitting, a cross-validation approach would partially mitigate it. In the future, we propose to create an interpretable approach using the dense embeddings themselves. Thirdly, we have not integrated demographic data from EHR with vitals and text data. Effectively, we propose to combine such datasets, along with the text and vitals based features for improving the predictions further. Lately, a major concern has been the failure of AI pipelines at bedside. We essentially plan to deploy our pipeline in a real-world setting for retrospective validation. Also, to demonstrate the impact of our architecture on long-term patient outcomes, we would like to extend our study in cohorts based on different time stamps (48 hours, 72 hours) and using additional outcome data such as mortality and length of ICU stay.

In conclusion, clinician notes are the closest representation of a clinician’s thought process for decision-making. Hence, they contain rich contextual information which has been underutilized for predictive settings. Also vitals data contains a large chunk of information, a subset of which is disjoint to the contextual information contained in clinical notes. This leads to better prediction when both the modalities of data (physician notes and vitals data) are utilized in critical care settings. The technical novelty of our pipeline and interpretable predictions can be leveraged to yield predictions for other critical outcomes through a transfer learning mechanism. Finally, we believe that work enables decision making in a variety of settings including resource constrained ICUs because of broader availability of clinician notes and vitals data.

## Data Availability

Data is available on reasonable request to the corresponding author.

## ACKNOWLEDGEMENTS

We acknowledge the support from the Center of Excellence in Healthcare at IIIT-Delhi.

## CONTRIBUTORSHIP STATEMENT

Conceptualization: R.P., T.S., P.M.

Cohort Building: A.B., P.S, A.N, S.P.

Methodology: R.P., A.B., S.P., P.S., R.S., P.M., T.S.

Visualization: R.P., S.P., R.S.

Clinical Inference: A.A., A.N., P.M., T.S., A.K.

Project administration: T.S., P.M, R.P.

Supervision: T.S., P.M.

Writing & Editing - R.P., S.P., A.B., A.A., P.M., R.L., T.S.

All authors read and approved the final paper.

## REFERENCES

1 Adams R, Henry KE, Sridharan A, et al. Prospective, multi-site study of patient outcomes after implementation of the TREWS machine learning-based early warning system for sepsis. Nat Med 2022;28:1455–60. doi:10.1038/s41591-022-01894-0

2 Ehwerhemuepha L, Heyming T, Marano R, et al. Development and validation of an early warning tool for sepsis and decompensation in children during emergency department triage. Sci Rep 2021;11:8578. doi:10.1038/s41598-021-87595-z

3 Kennedy JN, Rudd KE. A sepsis early warning system is associated with improved patient outcomes. Cell Rep Med 2022;3:100746. doi:10.1016/j.xcrm.2022.100746

4 Henry KE, Adams R, Parent C, et al. Factors driving provider adoption of the TREWS machine learning-based early warning system and its effects on sepsis treatment timing. Nat Med 2022;28:1447–54. doi:10.1038/s41591-022-01895-z

5 Hammoud I, Ramakrishnan I, Henry M, et al. Multimodal Early Septic Shock Prediction Model using Lasso Regression with Decaying Response. In: 2020 IEEE International Conference on Healthcare Informatics (ICHI). 2020. 1–3. doi:10.1109/ICHI48887.2020.9374377

6 Amrollahi F, Shashikumar SP, Razmi F, et al. Contextual Embeddings from Clinical Notes Improves Prediction of Sepsis. AMIA Annu Symp Proc 2021;2020:197–202.

7 Goh KH, Wang L, Yeow AYK, et al. Artificial intelligence in sepsis early prediction and diagnosis using unstructured data in healthcare. Nat Commun 2021;12:711. doi:10.1038/s41467-021-20910-4

8 Koch E, Lovett S, Nghiem T, et al. Shock index in the emergency department: utility and limitations. Open Access Emerg Med OAEM 2019;11:179–99. doi:10.2147/OAEM.S178358

9 Cheng T-H, Sie Y-D, Hsu K-H, et al. Shock Index: A Simple and Effective Clinical Adjunct in Predicting 60-Day Mortality in Advanced Cancer Patients at the Emergency Department. Int J Environ Res Public Health 2020;17:4904. doi:10.3390/ijerph17134904

10 Maheshwari K, Nathanson BH, Munson SH, et al. Abnormal shock index exposure and clinical outcomes among critically ill patients: A retrospective cohort analysis. J Crit Care 2020;57:5–12. doi:10.1016/j.jcrc.2020.01.024

11 Huang B, Yang Y, Zhu J, et al. Usefulness of the Admission Shock Index for Predicting Short-Term Outcomes in Patients With ST-Segment Elevation Myocardial Infarction. Am J Cardiol 2014;114:1315–21. doi:10.1016/j.amjcard.2014.07.062

12 Wc S. Temporal physiologic patterns of shock and circulatory dysfunction based on early descriptions by invasive and noninvasive monitoring. New Horiz Baltim Md 1996;4.https://pubmed.ncbi.nlm.nih.gov/8774804/ (accessed 9 Oct 2022).

13 Clinical pathology of the shock syndromes - PubMed. https://pubmed.ncbi.nlm.nih.gov/21769211/ (accessed 9 Oct 2022).

14 Assessment of Clinical Criteria for Sepsis: For the Third International Consensus Definitions for Sepsis and Septic Shock (Sepsis-3) | Critical Care Medicine | JAMA | JAMA Network. https://jamanetwork.com/journals/jama/fullarticle/2492875 (accessed 9 Oct 2022).

15 Predicting Hemodynamic Shock from Thermal Images using Machine Learning | Scientific Reports. https://www.nature.com/articles/s41598-018-36586-8 (accessed 15 Dec 2022).

16 Generalized Prediction of Hemodynamic Shock in Intensive Care Units | medRxiv. https://www.medrxiv.org/content/10.1101/2021.01.07.21249121v4 (accessed 15 Dec 2022).

17 Desautels T, Calvert J, Hoffman J, et al. Prediction of Sepsis in the Intensive Care Unit With Minimal Electronic Health Record Data: A Machine Learning Approach. JMIR Med Inform 2016;4:e28. doi:10.2196/medinform.5909

18 Gultepe E, Green JP, Nguyen H, et al. From vital signs to clinical outcomes for patients with sepsis: a machine learning basis for a clinical decision support system. J Am Med Inform Assoc JAMIA 2014;21:315–25. doi:10.1136/amiajnl-2013-001815

19 Potes C, Conroy B, Xu-Wilson M, et al. A clinical prediction model to identify patients at high risk of hemodynamic instability in the pediatric intensive care unit. Crit Care Lond Engl 2017;21:282. doi:10.1186/s13054-017-1874-z

20 Johnson AEW, Pollard TJ, Shen L, et al. MIMIC-III, a freely accessible critical care database. Sci Data 2016;3:160035. doi:10.1038/sdata.2016.35

21 Ramesh S, Tiwari A, Choubey P, et al. BERT based Transformers lead the way in Extraction of Health Information from Social Media. In: Proceedings of the Sixth Social Media Mining for Health (#SMM4H) Workshop and Shared Task. Mexico City, Mexico: : Association for Computational Linguistics 2021. 33–8. doi:10.18653/v1/2021.smm4h-1.5

22 Kormilitzin A, Vaci N, Liu Q, et al. Med7: A transferable clinical natural language processing model for electronic health records. Artif Intell Med 2021;118:102086. doi:10.1016/j.artmed.2021.102086

23 Mikolov T, Chen K, Corrado G, et al. Efficient Estimation of Word Representations in Vector Space. ArXiv13013781 Cs Published Online First: 6 September 2013.http://arxiv.org/abs/1301.3781 (accessed 27 Oct 2021).

24 Song X, Salcianu A, Song Y, et al. Fast WordPiece Tokenization. 2021. doi:10.48550/arXiv.2012.15524

25 Lee J, Yoon W, Kim S, et al. BioBERT: a pre-trained biomedical language representation model for biomedical text mining. Bioinformatics 2019;:btz682. doi:10.1093/bioinformatics/btz682

26 Alsentzer E, Murphy J, Boag W, et al. Publicly Available Clinical BERT Embeddings. In: Proceedings of the 2nd Clinical Natural Language Processing Workshop. Minneapolis, Minnesota, USA: : Association for Computational Linguistics 2019. 72–8. doi:10.18653/v1/W19-1909

27 Beltagy I, Peters ME, Cohan A. Longformer: The Long-Document Transformer. ArXiv200405150 Cs Published Online First: 2 December 2020.http://arxiv.org/abs/2004.05150 (accessed 27 Oct 2021).

28 Mulyar A, Schumacher E, Rouhizadeh M, et al. Phenotyping of Clinical Notes with Improved Document Classification Models Using Contextualized Neural Language Models. ArXiv191013664 Cs Published Online First: 16 September 2020.http://arxiv.org/abs/1910.13664 (accessed 27 Oct 2021).

29 Pappagari R, Zelasko P, Villalba J, et al. Hierarchical Transformers for Long Document Classification. ArXiv191010781 Cs Stat Published Online First: 23 October 2019.http://arxiv.org/abs/1910.10781 (accessed 27 Oct 2021).

30 Le QV, Mikolov T. Distributed Representations of Sentences and Documents. 2014. doi:10.48550/arXiv.1405.4053

31 Christ M, Braun N, Neuffer J, et al. Time Series FeatuRe Extraction on basis of Scalable Hypothesis tests (tsfresh –A Python package). Neurocomputing 2018;307. doi:10.1016/j.neucom.2018.03.067

32 Geurts P, Ernst D, Wehenkel L. Extremely randomized trees. Mach Learn 2006;63:3–42. doi:10.1007/s10994-006-6226-1

33 Pedregosa F, Varoquaux G, Gramfort A, et al. Scikit-learn: Machine Learning in Python. ArXiv12010490 Cs Published Online First: 5 June 2018.http://arxiv.org/abs/1201.0490 (accessed 27 Oct 2021).

34 Chawla NV, Bowyer KW, Hall LO, et al. SMOTE: Synthetic Minority Over-sampling Technique. J Artif Intell Res 2002;16:321–57. doi:10.1613/jair.953

35 Breiman L. Random Forests. Mach Learn 2001;45:5–32. doi:10.1023/A:1010933404324

36 Friedman JH. Stochastic gradient boosting. Comput Stat Data Anal 2002;38:367–78. doi:10.1016/S0167-9473(01)00065-2

37 Freund Y, Schapire RE. A Decision-Theoretic Generalization of On-Line Learning and an Application to Boosting. J Comput Syst Sci 1997;55:119–39. doi:10.1006/jcss.1997.1504

38 Chen T, Guestrin C. XGBoost: A Scalable Tree Boosting System. Proc 22nd ACM SIGKDD Int Conf Knowl Discov Data Min 2016;:785–94. doi:10.1145/2939672.2939785

39 Wright RE. Logistic regression. In: Reading and understanding multivariate statistics. Washington, DC, US:: American Psychological Association 1995. 217–44.

40 Lundberg SM, Lee S-I. A Unified Approach to Interpreting Model Predictions. In: Guyon I, Luxburg UV, Bengio S, et al., eds. Advances in Neural Information Processing Systems. Curran Associates, Inc. 2017. https://proceedings.neurips.cc/paper/2017/file/8a20a8621978632d76c43dfd28b67767-Paper.pdf

41 Wolf T, Debut L, Sanh V, et al. Transformers: State-of-the-Art Natural Language Processing. In: Proceedings of the 2020 Conference on Empirical Methods in Natural Language Processing: System Demonstrations. Online: : Association for Computational Linguistics 2020. 38–45. doi:10.18653/v1/2020.emnlp-demos.6

42 Surviving Sepsis Campaign: International Guidelines for Mana… : Critical Care Medicine. https://journals.lww.com/ccmjournal/fulltext/2021/11000/surviving_sepsis_campaign internat ional.21.aspx (accessed 9 Oct 2022).

43 Liu R, Greenstein JL, Sarma SV, et al. Natural Language Processing of Clinical Notes for Improved Early Prediction of Septic Shock in the ICU. In: 2019 41st Annual International Conference of the IEEE Engineering in Medicine and Biology Society (EMBC). Berlin, Germany: : IEEE 2019. 6103–8. doi:10.1109/EMBC.2019.8857819

44 Culliton P, Levinson M, Ehresman A, et al. Predicting Severe Sepsis Using Text from the Electronic Health Record. ArXiv171111536 Cs Published Online First: 30 November 2017.http://arxiv.org/abs/1711.11536 (accessed 12 May 2022).

