## Supplementary File for "*ShockModes:* A Multimodal Model for Prognosticating Intensive Care Outcomes from Physician Notes and Vitals"

### Supplementary Information

TITLE: Chief Complaint: pericardial effusion; shortness of breath HPI: 79 yo female with h/o COPD on home O2 DRUG , HTN, HLP, DM, osteoporosis, morbid obesity who presented to an OSH with worsening SOB. She had been treated with Z-pack DRUG without benefit. She continued to have worsening SOB throughout her admission there. She was started on a prednisone DRUG taper DOSAGE for a COPD exacerbation. She had an echocardiogram initially which showed a pericardial effusion, which was fairly small without any evidence of tamponade physiology, but she then had a repeat echo cardiogram on [**11-5**] with which showed a large anterior and posterior pericardial effusion which measured 17 mm in diastole and 21 mm in systole. She was then transferred for further evaluation and potential pericardiocentesis. Currently, she feels slightly better than before. She would just like to have something done with this fluid so that she can go home soon. She is also concerned about the origin of the fluid. On review of systems, she denies any prior history of stroke, TIA, deep venous thrombosis, pulmonary embolism, bleeding at the time of surgery, myalgias, joint pains, cough, hemoptysis, black stools or red stools. She denies recent fevers, chills or rigors. She denies exertional buttock or calf pain. All of the other review of systems were negative. Cardiac review of systems is notable for absence of chest pain, paroxysmal nocturnal dyspnea, orthopnea, palpitations, syncope or presyncope. She does note some dyspnea on exertion and ankle edema. Patient admitted from: Transfer from other hospital History obtained from Patient Allergies: Penicillins DRUG Rash; Sulfa (Sulfonamide Antibiotics DRUG ) Rash; Last dose of Antibiotics DRUG : Infusions: Other ICU medications: Other medications: MEDICATIONS AT HOME: Lasix DRUG 40 mg STRENGTH daily FREQUENCY Antivert DRUG 25 mg STRENGTH TID FREQUENCY Lipitor DRUG 10 mg STRENGTH daily FREQUENCY Prilosec DRUG 20 mg STRENGTH daily FREQUENCY Synthroid DRUG 125 mcg STRENGTH daily FREQUENCY Xalatan DRUG 0.005 STRENGTH % each eye ROUTE qhs FREQUENCY Advair DRUG diskus FORM 500/50 mcg STRENGTH [**Hospital1 7**] Nasonex DRUG 1 DOSAGE squirt FORM in each nose [**Hospital1 7**] Avandamet DRUG 25/250 mg STRENGTH [**Hospital1 7**] Albuterol DRUG 2.5 mg STRENGTH q2H nebulized FREQUENCY PRN FREQUENCY Claritin DRUG 10 mg STRENGTH QHS FREQUENCY Fosamax DRUG 70 mg STRENGTH daily FREQUENCY Hydroxyzine DRUG . MEDICATIONS AT TRANSFER: Prednisone DRUG 50 mg STRENGTH daily FREQUENCY - taper DOSAGE Metoprolol tartrate DRUG 12.5 mg STRENGTH [**Hospital1 7**] Synthroid DRUG 125 mcg STRENGTH daily FREQUENCY Prevacid DRUG 30 mg STRENGTH [**Hospital1 7**] Nystatin DRUG powder FORM daily FREQUENCY Lasix DRUG 40 mg STRENGTH daily FREQUENCY Lipitor DRUG 10 mg STRENGTH daily FREQUENCY Xalatan DRUG 0.005% STRENGTH QHS FREQUENCY each eye ROUTE Nasonex DRUG [**Hospital1 7**] Claritin DRUG 10 mg STRENGTH daily FREQUENCY Heparin DRUG sc ROUTE Tylenol DRUG PRN FREQUENCY Lorazepam DRUG 0.5 mg STRENGTH Q6H FREQUENCY PRN Antivert DRUG 25 mg STRENGTH PO ROUTE Q6H PRN FREQUENCY dizziness Past medical history: Family history: Social History: 1. CARDIAC RISK FACTORS: (+)Diabetes, (+)Dyslipidemia, (+)Hypertension 2. CARDIAC HISTORY: -CABG: -PERCUTANEOUS CORONARY INTERVENTIONS: -PACING/ICD: 3. OTHER PAST MEDICAL HISTORY: COPD on home O2 DRUG Diverticulitis Morbid Obesity Hypothyroidism Chronic LBP Chronic Headaches Glaucoma No family history of early

*Supplementary Fig 1: A sample of physician notes where clinical Named Entity Recognition (NER) has been performed using Med7. Different entities such as Drugs, Dosage, Frequency, Strength have been contrasted in the text with different colors. We have used only the Drug entity for modeling the instances of Shock/Non-Schock.*

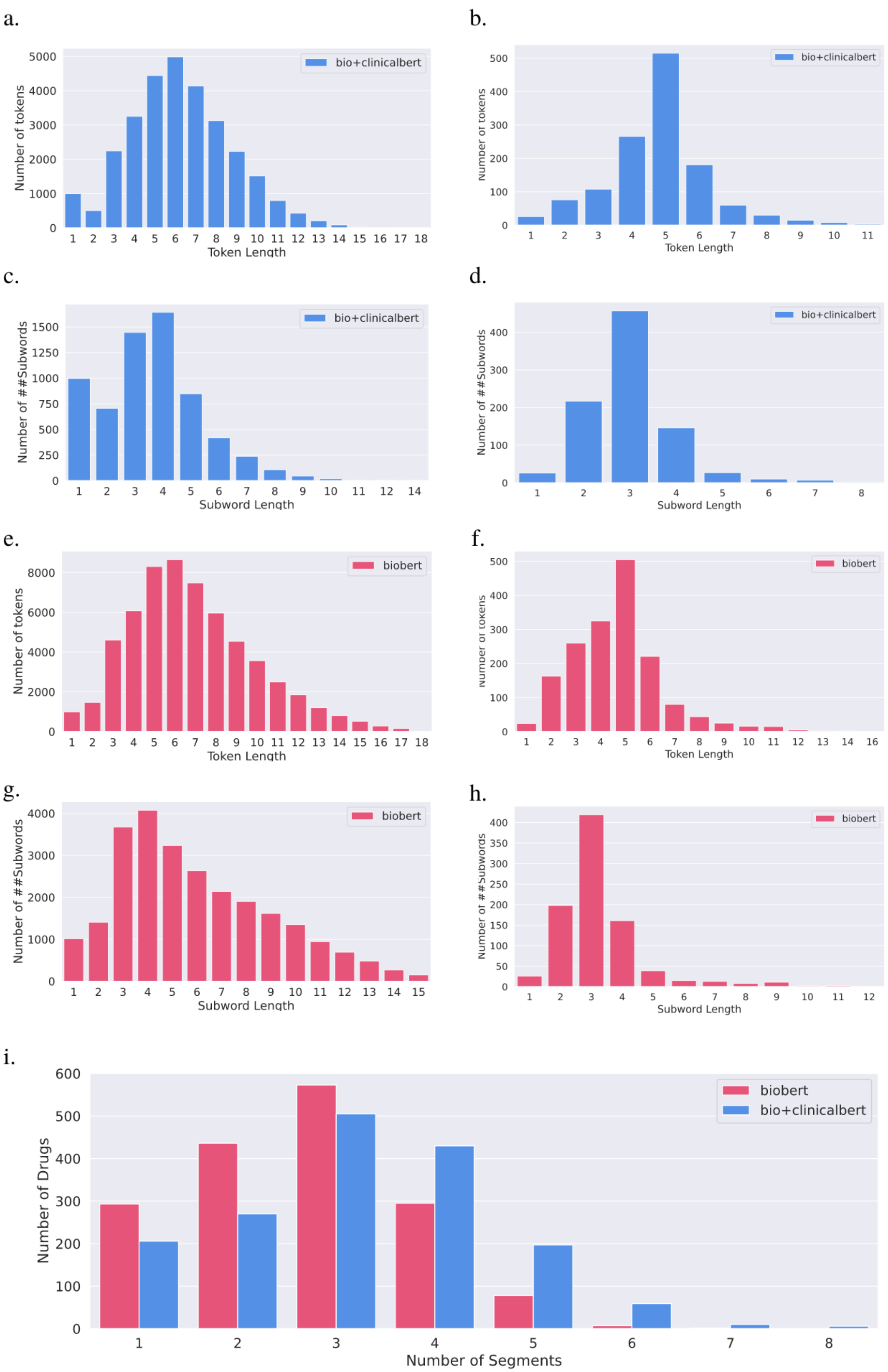

a.
bio+clinicalbert
Number of tokens
Token Length
b.
bio+clinicalbert
Number of tokens
Token Length
c.
bio+clinicalbert
Number of ##Subwords
Subword Length
d.
bio+clinicalbert
Number of ##Subwords
Subword Length
e.
biobert
Number of tokens
Token Length
f.
biobert
Number of tokens
Token Length
g.
biobert
Number of ##Subwords
Subword Length
h.
biobert
Number of ##Subwords
Subword Length
i.
biobert
bio+clinicalbert
Number of Drugs
Number of Segments

*Supplementary Fig2: Visualizations for Bio-Bert and Bio+Clinical-Bert vocabulary, and tokens generated from physician notes a,b) token length distribution for Bio+Clinical-Bert vocabulary and tokens generated from physician notes, c,d) subword length (length of tokens with ## Wordpiece representation without ## characters) distribution for Bio+Clinical-Bert vocabulary and generated tokens, e,f) token length distribution for Bio-Bert vocabulary and generated tokens, g,h) subword length distribution for Bio-Bert vocabulary and generated tokens, i) Comparison between segment length (number of words/subwords used to represent a word) distribution for tokens generated using physician notes by Bio-Bert and Bio+Clinical-Bert.*

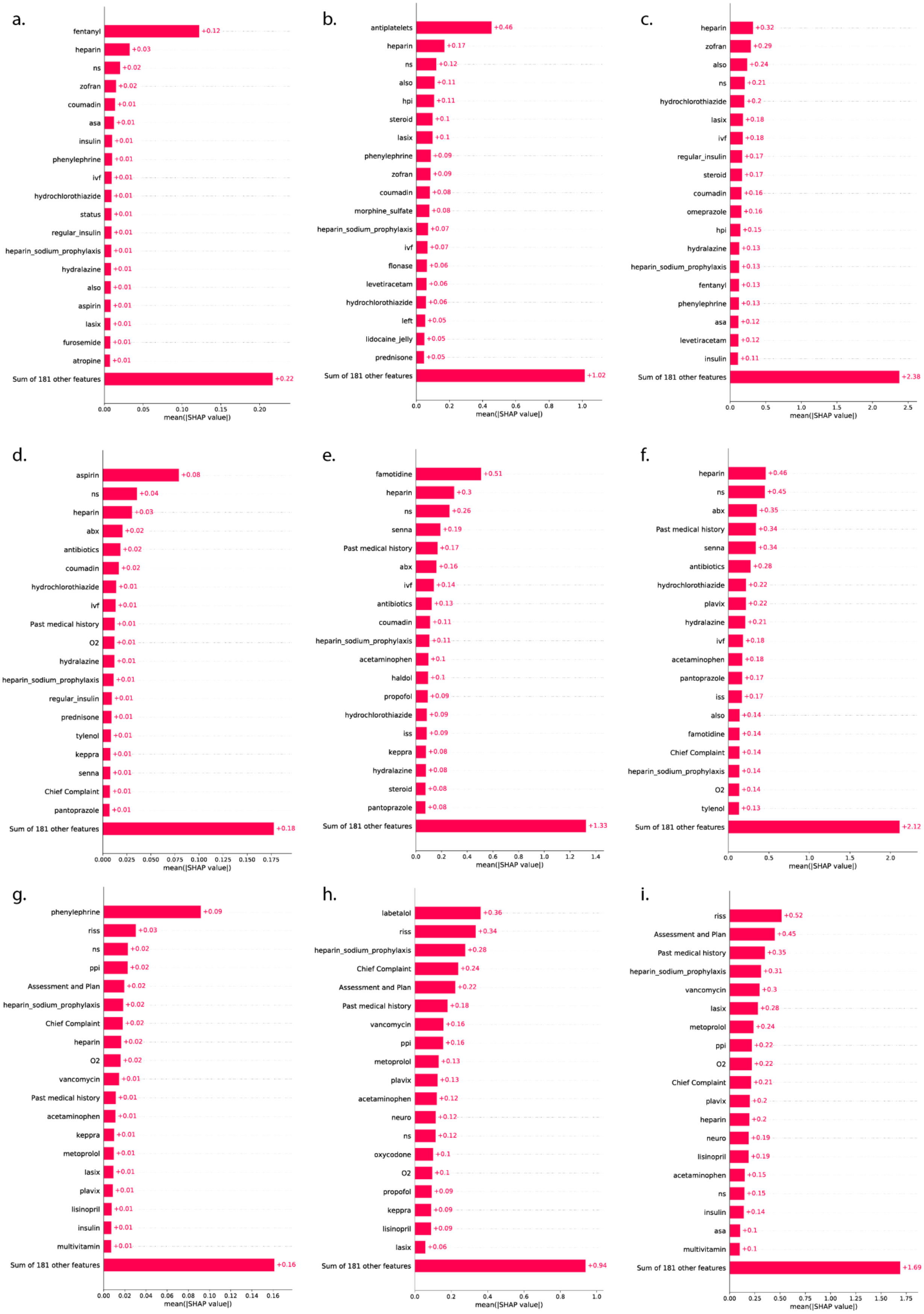

a.
fentanyl +0.12
heparin +0.03
ns +0.02
zofran +0.02
coumadin +0.01
asa +0.01
insulin +0.01
phenylephrine +0.01
ivf +0.01
hydrochlorothiazide +0.01
status +0.01
regular_insulin +0.01
heparin_sodium_prophylaxis +0.01
hydralazine +0.01
also +0.01
aspirin +0.01
lasix +0.01
furosemide +0.01
atropine +0.01
Sum of 181 other features +0.22
0.00 0.05 0.10 0.15 0.20
mean(|SHAP value|)
b.
antiplatelets +0.46
heparin +0.17
ns +0.12
also +0.11
hpi +0.11
steroid +0.1
lasix +0.1
phenylephrine +0.09
zofran +0.09
coumadin +0.08
morphine_sulfate +0.08
heparin_sodium_prophylaxis +0.07
ivf +0.07
flonase +0.06
levetiracetam +0.06
hydrochlorothiazide +0.06
left +0.05
lidocaine_jelly +0.05
prednisone +0.05
Sum of 181 other features +1.02
0.0 0.2 0.4 0.6 0.8 1.0
mean(|SHAP value|)
c.
heparin +0.32
zofran +0.29
also +0.24
ns +0.21
hydrochlorothiazide +0.2
lasix +0.18
ivf +0.18
regular_insulin +0.17
steroid +0.17
coumadin +0.16
omeprazole +0.16
hpi +0.15
hydralazine +0.13
heparin_sodium_prophylaxis +0.13
fentanyl +0.13
phenylephrine +0.13
asa +0.12
levetiracetam +0.12
insulin +0.11
Sum of 181 other features +2.38
0.0 0.5 1.0 1.5 2.0 2.5
mean(|SHAP value|)
d.
aspirin +0.08
ns +0.04
heparin +0.03
abx +0.02
antibiotics +0.02
coumadin +0.02
hydrochlorothiazide +0.01
ivf +0.01
Past medical history +0.01
O2 +0.01
hydralazine +0.01
heparin_sodium_prophylaxis +0.01
regular_insulin +0.01
prednisone +0.01
tylenol +0.01
keppra +0.01
senna +0.01
Chief Complaint +0.01
pantoprazole +0.01
Sum of 181 other features +0.18
0.000 0.025 0.050 0.075 0.100 0.125 0.150 0.175
mean(|SHAP value|)
e.
famotidine +0.51
heparin +0.3
ns +0.26
senna +0.19
Past medical history +0.17
abx +0.16
ivf +0.14
antibiotics +0.13
coumadin +0.11
heparin_sodium_prophylaxis +0.11
acetaminophen +0.1
haldol +0.1
propofol +0.09
hydrochlorothiazide +0.09
iss +0.09
keppra +0.08
hydralazine +0.08
steroid +0.08
pantoprazole +0.08
Sum of 181 other features +1.33
0.0 0.2 0.4 0.6 0.8 1.0 1.2 1.4
mean(|SHAP value|)
f.
heparin +0.46
ns +0.45
abx +0.35
Past medical history +0.34
senna +0.34
antibiotics +0.28
hydrochlorothiazide +0.22
plavix +0.22
hydralazine +0.21
ivf +0.18
acetaminophen +0.18
pantoprazole +0.17
iss +0.17
also +0.14
famotidine +0.14
Chief Complaint +0.14
heparin_sodium_prophylaxis +0.14
O2 +0.14
tylenol +0.13
Sum of 181 other features +2.12
0.0 0.5 1.0 1.5 2.0
mean(|SHAP value|)
g.
phenylephrine +0.09
riss +0.03
ns +0.02
ppi +0.02
Assessment and Plan +0.02
heparin_sodium_prophylaxis +0.02
Chief Complaint +0.02
heparin +0.02
O2 +0.02
vancomycin +0.01
Past medical history +0.01
acetaminophen +0.01
keppra +0.01
metoprolol +0.01
lasix +0.01
plavix +0.01
lisinopril +0.01
insulin +0.01
multivitamin +0.01
Sum of 181 other features +0.16
0.00 0.02 0.04 0.06 0.08 0.10 0.12 0.14 0.16
mean(|SHAP value|)
h.
labetalol +0.36
riss +0.34
heparin_sodium_prophylaxis +0.28
Chief Complaint +0.24
Assessment and Plan +0.22
Past medical history +0.18
vancomycin +0.16
ppi +0.16
metoprolol +0.13
plavix +0.13
acetaminophen +0.12
neuro +0.12
ns +0.12
oxycodone +0.1
O2 +0.1
propofol +0.09
keppra +0.09
lisinopril +0.09
lasix +0.06
Sum of 181 other features +0.94
0.0 0.2 0.4 0.6 0.8 1.0
mean(|SHAP value|)
i.
riss +0.52
Assessment and Plan +0.45
Past medical history +0.35
heparin_sodium_prophylaxis +0.31
vancomycin +0.3
lasix +0.28
metoprolol +0.24
ppi +0.22
O2 +0.22
Chief Complaint +0.21
plavix +0.2
heparin +0.2
neuro +0.19
lisinopril +0.19
acetaminophen +0.15
ns +0.15
insulin +0.14
asa +0.1
multivitamin +0.1
Sum of 181 other features +1.69
0.00 0.25 0.50 0.75 1.00 1.25 1.50 1.75
mean(|SHAP value|)

*Supplementary Fig3: a,b,c) Global features importance analysis with Word2vec as therapeutics embedding and Doc2vec as HOPI embedding. d,e,f) Global features importance analysis with BioClinical-BERT as therapeutics embedding and DocBERT as HOPI embedding. g,h,i) Global features importance analysis with Bio-BERT as therapeutics embedding and DocBERT as HOPI embedding.*

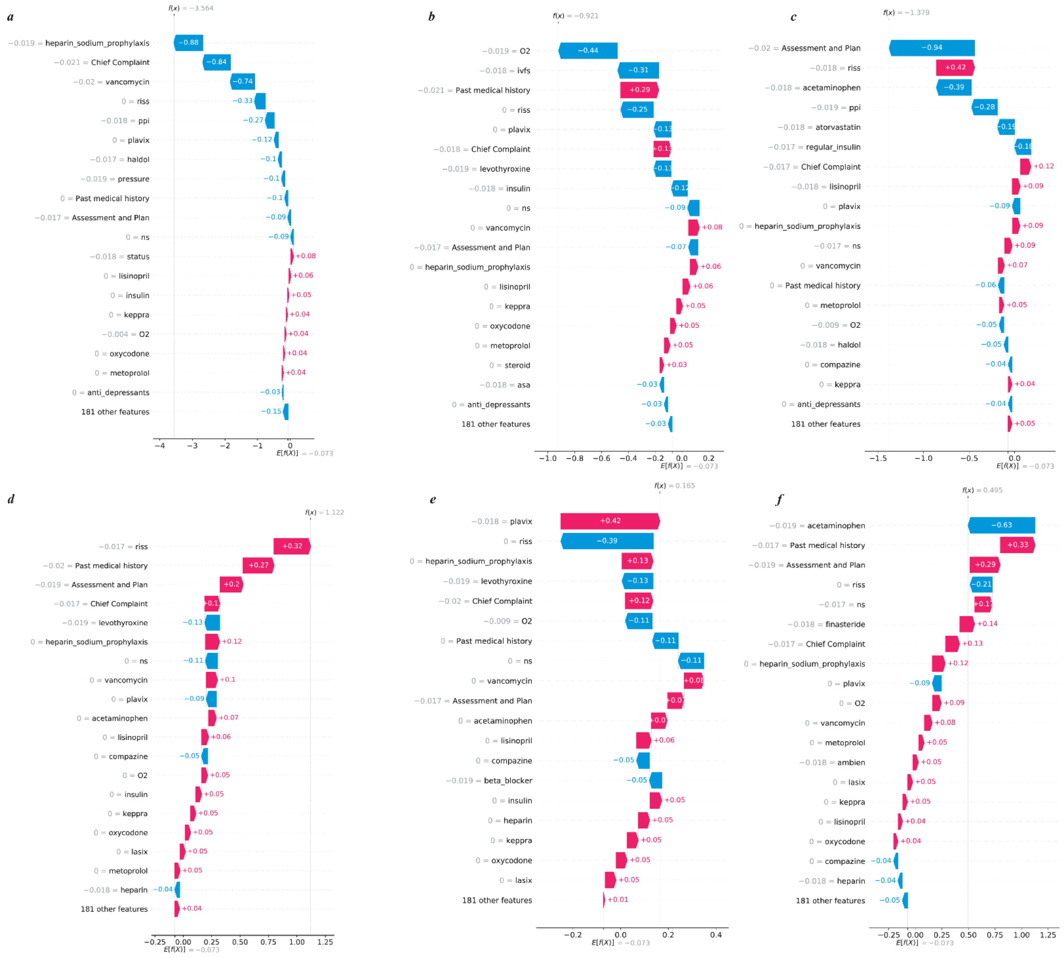


*Supplementary Fig4: Validation of few instances of patients where the model outcome was accurate a,b,c) Waterfall plot for interpreting the model outcome at an individual level where the model outcome was Non-shock and true label also depicted Non-shock, d,e,f) Waterfall plot for interpreting the model outcome at an individual level where the model outcome was Shock and true label also depicted Shock.*

| Variables | Median (IQR) | | Statistical Test |
|---|---|---|---|
| | Abnormal Shock Index | Normal Shock Index | p-value |
| Age | 68.99 (19.37) | 69.28 (22.17) | 0.927 (w)* |
| Length of stay(LOS) | 4.25 (7.45) | 4.95 (7.99) | 0.180 (w)* |
| Heart Rate, per min | 72 (10.50) | 68 (10.00) | $8.14 \times 10^{-07}$ (w)* |
| Pulse Rate, per min | 78 (10.00) | 67 (11.00) | $3.89 \times 10^{-06}$ (w)* |
| Respiratory rate, per min | 19 (4.00) | 18 (4.66) | 0.101 (w)* |
| Oxygen Saturation | 97 (3.08) | 97 (3.00) | 0.886 (w)* |
| Gender (Female %) | 39.08% | 38.87% | 0.903 (c)* |

*Supplementary Table 1. Textual Cohort characteristics. Values are median (IQR) unless indicated, *signifies Wilcoxon (W) Ranksum test (non-parametric) or Student's t-test (t) (parametric) which were used after testing for the normality assumption. c - Chi-squared test of proportions.*

| Models | Parameters |
|---|---|
| Word2Vec | vector_size=100, window=5, min_count=1, sg=1, epochs=5,<br>Default parameters from gensim.models.Word2Vec |
| Doc2Vec | dm=1, min_alpa=0.00025, min_count=1, epochs=5,<br>Default parameters from gensim.models.doc2vec.Doc2Vec |
| Random Forest | n_estimators(trees)=100,<br>Default parameters from sklearn.ensemble.RandomForestClassifier |
| GB Boost | learning_rate=0.1,<br>Default parameters from sklearn.ensemble.GradientBoostingClassifier |

| | |
|---|---|
| XGBoost | n_estimators(trees)=50,<br>Default parameters from xgboost.XGBClassifier |
| AdaBoost | n_estimators(tress)=50, learning_rate=0.1,<br>Default parameters from sklearn.ensemble.AdaBoostClassifier |
| BioBERT | Default parameters from Huggingface |
| BioClinicalBERT | Default parameters from Huggingface |

*Supplementary Table 2: Model and respective parameters used for training purpose.*

| RESOURCE | SOURCE | IDENTIFIER |
|---|---|---|
| **Software and Algorithms** | | |
| Anaconda (version 4.5.12) | anaconda.com | https://www.anaconda.com/ |
| Python (version 3.6.5) | python.org | https://www.python.org/downloads/release/python-365/ |
| gensim (version 3.8.1) | PyPI | https://pypi.org/project/gensim/3.8.1/ |
| numpy (version 1.19.0) | PyPI | https://pypi.org/project/numpy/1.19.0/ |
| pandas (version 1.0.5) | PyPI | https://pypi.org/project/pandas/1.0.5/ |
| nltk (version 3.4.5) | PyPI | https://pypi.org/project/nltk/3.4.5/ |
| strsimpy (version 0.1.9) | PyPI | https://pypi.org/project/strsimpy/0.1.9/ |
| spacy (version 2.3.5) | PyPI | https://pypi.org/project/spacy/2.3.5/ |
| sklearn (version 0.23.2) | PyPI | https://pypi.org/project/scikit-learn/0.23.2/ |
| en_core_med7_lg (version 0.0.3) | PyPI | https://med7.s3.eu-west-2.amazonaws.com/en_core_med7_lg.tar.gz |
| plotly | PyPI | https://pypi.org/project/plotly/4.14.3/ |
| imbalanced-learn | PyPI | https://pypi.org/project/imbalanced-learn/0.7.0/ |
| hugging face (version 0.0.1) | PyPI | https://pypi.org/project/huggingface/0.0.1/ |
| transformers (version 4.10.2) | PyPI | https://pypi.org/project/transformers/4.10.2/ |
| sentencepiece (version 0.1.96) | PyPI | https://pypi.org/project/sentencepiece/0.1.96/ |
| shap (version 0.38.1) | PyPI | https://pypi.org/project/shap/0.38.1/ |

*Supplementary Table 3: List of software and packages used for our study with their sources and identifiers for the reproducibility of this study.*

| Model | Accuracy | Precision | Recall | F1 Score | AUC-ROC |
|---|---|---|---|---|---|
| **Random Forest** | p<.05 | p<.05 | p<.05 | p>.05 | p<.05 |
| **Gradient boosting** | p<.05 | p=.05 | p<.05 | p<.05 | p<.05 |
| **Adaptive Boosting** | p<.05 | p<.05 | p<.05 | p<.05 | p<.05 |
| **Extreme Gradient Boosting** | p>.05 | p<.05 | p>.05 | p<.05 | p=.05 |

*Supplementary Table 4: Welch's t-test results on the performance algorithms for the importance of History of Present Illness as a feature (Comparison of models with and without diagnosis). The mean accuracy and recall of all algorithms were found to be significantly different from each other, whereas for precision, F1-score and AUC-ROC score there was one value for each metric which was not statistically significant.*

| **Therapeutics Only as Features** | | | | | | |
|---|---|---|---|---|---|---|
| **Model** | **Accuracy** | **Precision** | **Recall** | **F1 Score** | **AUC-ROC** | **Specificity** |
| **BioBERT** | | | | | | |
| **Logistic Regression** | 54.02 ± 0.0108 | 0.78 ± 0.0107 | 0.54 ± 0.0108 | 0.59 ± 0.0102 | 0.68 ± 0.0176 | 0.50 ± 0.0120 |
| **Random Forest** | 79.49 ± 0.79 | 0.71 ± 0.0139 | 0.79 ± 0.0079 | 0.74 ± 0.0103 | 0.59 ± 0.0146 | 0.96 ± 0.0039 |
| **Gradient boosting** | 76.46 ± 0.79 | 0.73 ± 0.0107 | 0.76 ± 0.0079 | 0.74 ± 0.0093 | 0.63 ± 0.0139 | 0.90 ± 0.0064 |
| **Adaptive Boosting** | 79.68 ± 0.80 | 0.75 ± 0.0111 | 0.79 ± 0.0080 | 0.76 ± 0.0098 | 0.67 ± 0.0151 | 0.94 ± 0.0052 |
| **Extreme Gradient Boosting** | 78.78 ± 0.77 | 0.73 ± 0.0115 | 0.79 ± 0.0077 | 0.75 ± 0.0098 | 0.60 ± 0.0151 | 0.94 ± 0.0048 |
| **Therapeutic + History of Present Illness as Features** | | | | | | |
| **BioBERT** | | | | | | |

| | | | | | | |
|---|---|---|---|---|---|---|
| **Logistic regression** | 53.53 ± 0.0108 | 0.78 ± 0.0107 | 0.54 ± 0.0108 | 0.58 ± 0.0101 | 0.62 ± 0.0157 | 0.49 ± 0.0118 |
| **Random Forest** | **80.9 ± 0.72** | 0.67 ± 0.0116 | **0.81 ± 0.0072** | 0.73 ± 0.0099 | 0.60 ± 0.0147 | 0.99 ± 0.0021 |
| **Gradient boosting** | 79.36 ± 0.79 | 0.75 ± 0.0111 | 0.79 ± 0.0079 | 0.76 ± 0.0096 | **0.68 ± 0.0139** | 0.93 ± 0.0053 |
| **Adaptive Boosting** | 77.94 ± 0.80 | 0.73 ± 0.0112 | 0.78 ± 0.0080 | 0.74 ± 0.0096 | 0.65 ± 0.0151 | 0.93 ± 0.0054 |
| **Extreme Gradient Boosting** | 78.94 ± 0.83 | 0.76 ± 0.0107 | 0.79 ± 0.0083 | **0.77 ± 0.0096** | 0.61 ± 0.0157 | 0.92 ± 0.0059 |

*Supplementary Table 5. Analysis of the importance of History of Present Illness as features for the models with a margin error of one standard-deviation from the mean. Random Sampling(N=100) of the test dataset, with Bootstrap iterations of 100 has been recorded for the mean value.*

| Model | Accuracy | Precision | Recall | F1 Score | AUC-ROC |
|---|---|---|---|---|---|
| **Random Forest** | 70.33 + 0.91 | 0.70 + 0.0119 | 0.70 + 0.0091 | 0.70 + 0.0100 | 0.55 + 0.0135 |
| **Gradient boosting** | **72.94 + 0.85** | 0.69 + 0.0117 | **0.73 + 0.0085** | 0.71 + 0.0099 | 0.56 + 0.0159 |
| **Adaptive Boosting** | 72.26 + 0.91 | **0.73 + 0.0104** | 0.72 + 0.0091 | **0.72 + 0.0093** | **0.57 + 0.0160** |
| **Extreme Gradient Boosting** | 70.67 + 0.93 | 0.70 + 0.0119 | 0.71 + 0.0093 | 0.70 + 0.0102 | 0.52 + 0.0112 |

*Supplementary Table 6. Results of Hemodynamic shock prediction using drug embeddings generated from BioBert and considering non-mean embeddings of History of Present Illness(HOPI) from DocBert on a 24-hour cohort of MIMIC III dataset with a margin error of one standard-deviation from the mean. Random Sampling(N=100) of the test dataset, with Bootstrap iterations of 100 has been recorded for the mean value.*

Local feature importances for each individual was also calculated to understand patient-specific interpretability. Figures 4c and 4d are illustrative of feature importance for a representative patient with Shapley values indicating the likelihood of normal(blue) or abnormal SI(red). For this patient, the presence of levothyroxine and heparin as features were indicative of normal status 24 hours later. Figure 4d shows a physician note for prediction of abnormal SI with RISS as a feature suggestive of downstream development of abnormal shock index. Validation of model performance based on interpretability for a few other instances of abnormal/normal SI have been supplemented (Supplementary Figure 4).

The explainability of models is a major concern in the healthcare domain.[62] We address this concern of explainability using SHAP. The bar plot(Figure 4a) demonstrates RISS, Heparin sodium prophylaxis, HOPI, and Vancomycin as the most important features in modeling. Clinician assessment of the patient included in chief complaint, assessment and plan, past medical history, are all indicative of the type of illness and severity. Whereas early therapeutic interventions are all indicative of the efforts to prevent future deterioration as a followup to the above assessment. Hence, fluids such as NS (Normal Saline), antibiotics such as Vancomycin, supplemental oxygen as supportive treatment and Propofol for sedation of mechanically ventilated patients, explain the critical illness of the patient and are indicative of future risk of development of shock compared to patients who are not receiving these therapies. Similarly, patients receiving RISS to treat hyperglycemia or heparin for venous thromboembolism prophylaxis represent those who are sicker, and have higher likelihood of deterioration into shock.
